# User profile, indications and relevance of MRI in public and private health facilities in Cameroon

**DOI:** 10.1101/2025.05.18.25327866

**Authors:** Guy Roger Pilo Ndibo, Boniface Moifo, Joshua Tambe Agbor, Jean Roger Moulion Tapouh, Fabrice Zobel Lekeumo Cheuyem

## Abstract

**Objectives:** The aim of this study was to evaluate the relevance of MRI indications in the cities of Yaoundé and Douala and identify the factors associated with its accessibility and appropriate use.

**Materials and Methods:** We conducted a descriptive and analytical cross-sectional study with prospective data collection. We included all patients who underwent MRI during the study period with their informed consent. The main variables studied were sociodemographic variables (age, sex, occupation, health insurance), MRI-related variables (relevance of the examination, part of the body examined, indication for the examination, quality of the person requesting the examination) and clinical variables (patient’s comorbidities). Logistic regression with bivariate and multivariate analyses was used to determine the factors associated with the appropriate use of MRI, using SPSS version 26 software. The statistical significance level was set at 0.05.

**Results:** Our population consisted of 337 individuals, 174 of whom were women (51.6%). The median age of the study population was 48 years, with a minimum of 01 years and a maximum of 87 years. Patients aged ≥ 60 years were the most represented (27.9%). Participants were covered by some form of insurance in 20.5% of cases. The main prescribers were medical specialists (78.9%). Cerebral MRI was the most frequently performed procedure (39.5%). The test was considered relevant in 78.3% of cases and was mostly indicated as a first-line treatment (56.1%). After multivariate analysis, only the search for vascular pathologies was associated with the relevance of MRI as an imaging test.

**Conclusion:** The use of magnetic resonance imaging in our context is relevant in the majority of cases, and its use in the search for vascular pathology plays an important role in the relevance of this examination. In the light of these results, it would seem important to improve the affordability of MRI throughout the country, to increase the relevance of MRI indications and to optimize the training of medical staff in the justification of imaging examinations in general and MRI in particular.

## Background

Medical imaging has experienced significant advancements with the advent of increasingly high-performing techniques, enhancing the diagnostic capabilities for pathologies that were previously difficult to access [1]. Magnetic Resonance Imaging (MRI) is one of these sophisticated techniques whose availability is progressively increasing, including in developing countries [2]. It allows for better tissue characterization without ionizing radiation. MRI is a non-invasive technique with very high safety and significant diagnostic efficacy in various pathologies [3].

Thanks to its superior soft tissue contrast, MRI enables much more precise imaging of the neural axis and large joints of the musculoskeletal system, where it has become a reference standard [4]. Since its invention, the scope and application of magnetic resonance imaging have considerably expanded and now encompass abdominopelvic and cardiac imaging with dynamic sequences [4]. A major advantage of MRI is its ability to produce high-quality images with better soft tissue resolution without using ionizing radiation. The magnet generates images based on the specific and unique magnetic properties of tissues, driven by the spin properties of hydrogen molecules [5].

As with any new technology, we are witnessing an increasing use of MRI, with a non-negligible proportion of unjustified and abusive use [6]. In developing countries like Cameroon, the cost of an MRI is 3 to 5 times the minimum wage, and the health coverage rate is only 6.46% of the population [7]. Therefore, the use of this technique should be as appropriate as possible. Furthermore, various conflicts of interest, the phenomenon of incentive bonuses for prescribing in certain private centers, and the living conditions of young doctors, among other factors, could increase inappropriate MRI requests, as is the case with other imaging modalities. Conversely, the high cost of this examination could be a barrier to access.

In the context of improving healthcare provision, some health facilities have been equipped with MRI. Despite this, MRI remains scarcely available and quite difficult for the general population to access due to its cost, given that almost the entire cost of care is borne by the patient and their family. Thus, this imaging modality, although high-performing, is still considered a luxury by both prescribers and patients, who sometimes prefer to resort to other modalities for economic reasons. Furthermore, inappropriate prescriptions and performance of MRI examinations are detrimental to patients. We therefore conducted this study with the aim to assess sociodemographic profile of MRI users, the indications, and the relevance of using MRI in the Cameroonian context.

## Methods

### Study design

This was a descriptive and analytical cross-sectional study with prospective data collection.

### Study sites, period, and duration

The study was conducted from November 2022 to July 2023 in four healthcare facilities (three public and one private) in the cities of Douala and Yaoundé, which had a functional MRI machine during the study period. These included Yaoundé General Hospital, Military Regional Hospital Number 1, Laquintinie Hospital of Douala, and La Cathédrale Medical Center of Yaoundé.

### Study population

The study population consisted of all patients coming for an MRI examination in one of the aforementioned healthcare facilities.

### Study participant and sampling

Any patient who has undergone an MRI and has freely consented to participate in the study. We used consecutive and exhaustive sampling of all eligible patients. According to Cochran’s formula and anticipating a non-response rate of 15%, we obtained a minimum sample size of 441 participants.

### Data collection tool and procedure

A data collection tool was developed for the purpose of this study. We collected data related to sociodemographic characteristics (age, sex, insurance), the examination request and the patient’s clinical profile (body part examined, clinical information, type of prescriber). At each study site, participants were approached and informed about the study. After obtaining their free and informed consent, they were interviewed.

### Variable

The patient’s clinical information was classified into pathology groups including traumatic pathology, vascular pathology, congenital pathology, connective tissue disease/autoimmune disease, degenerative pathology, inflammatory/infectious pathology, metabolic/endocrine pathology, benign neoplasm, malignant neoplasm, non-specific pathology, others. Examination requests were divided into two categories: those with adequate clinical details for a particular clinical situation (appropriate request) and those with inadequate clinical details or a lack of indication (inappropriate request). To determine the relevance of the request, the French Society of Radiology (SFR) guidelines for the appropriate use of medical imaging examinations were used as a reference.

### Statistical Analysis

The data were entered and analyzed using SPSS software version 26.0. Quantitative variables were expressed as counts and percentages. Quantitative variables were expressed as mean with standard deviation or median with interquartile range, depending on the data distribution. The search for factors associated with the appropriate use of MRI was performed using Chi-square and Fisher’s Exact tests. The crude and adjusted Odds Ratio was used the assessed the strength of the association between variables. Data were considered statistically significant for a *p-*value < 0.05 with a 95% confidence interval.

## Results

A total of 337 patients were included during the study period. The median age of patients was 48 (Q1-Q3=35-60) years, with extremes of 1 and 87 years. Patients aged over 60 were the most common (27.9%). There was almost equal representation of both sexes (sex ratio: male/female = 0.98) and 20.5% had health insurance cover (Table 1).

**Table 1.** Socio-demographic profile of patients included, in Yaoundé, Cameroon, 2022-2023 (n=337)

| Variable | Count (n) | Frequency (%) |
| --- | --- | --- |
| **Age group** |  |  |
| < 21 | 67 | 19,9 |
| 21-30 | 57 | 16,9 |
| 30-40 | 27 | 8,0 |
| 40-50 | 30 | 8,9 |
| 50-60 | 61 | 18,1 |
| 60+ | 95 | 28,2 |
| **Gender** |  |  |
| Male | 163 | 48,4 |
| Female | 174 | 51,6 |
| **Health insurance** |  |  |
| Yes | 69 | 20,5 |
| No | 268 | 79,5 |

MRI scans were mainly requested by specialists (Fig 2).

**Fig 2.**
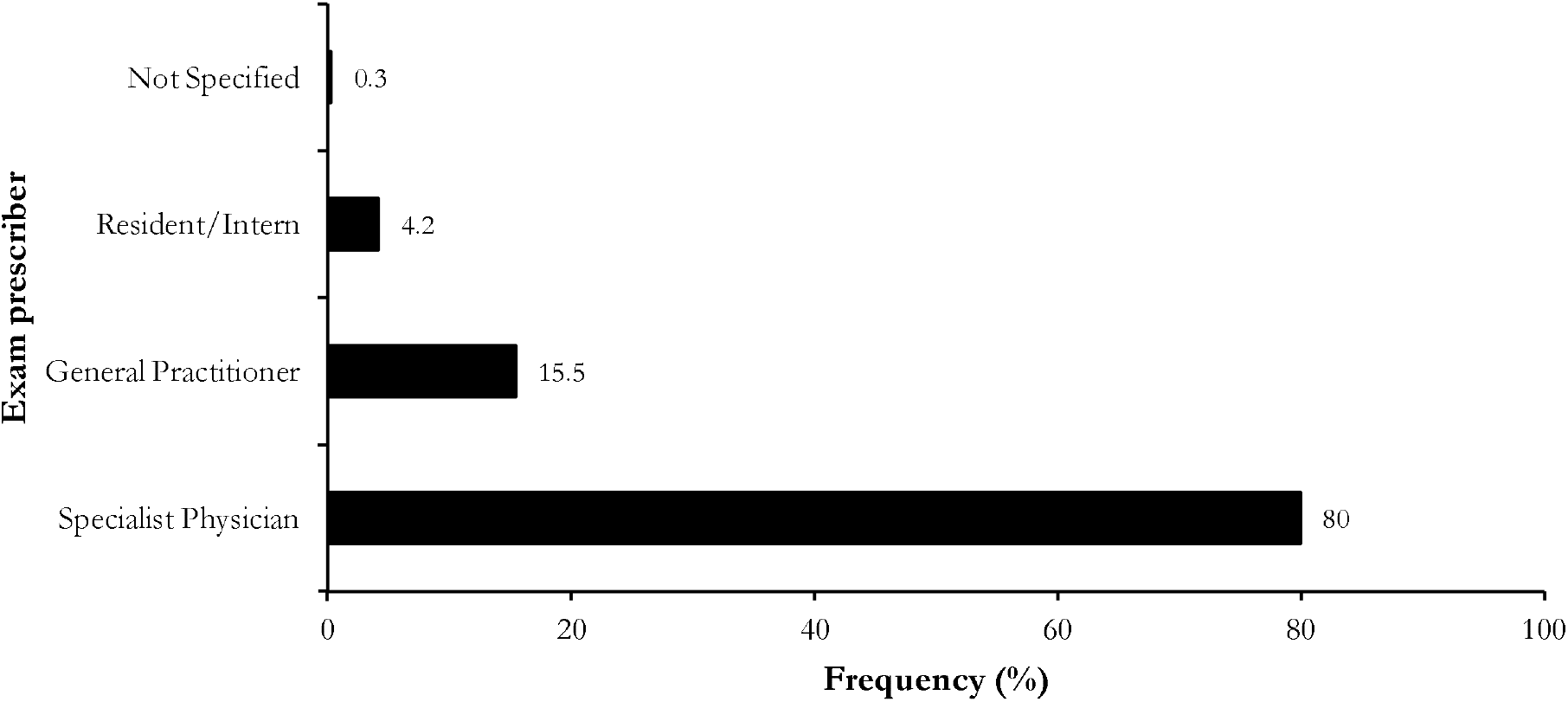
Status of the person ordering the MRI test in Yaoundé, Cameroon, 2022-2023.

The brain and lumbosacral spine accounted for 60.3% of the examinations requested (Fig 3).

**Fig 3.**
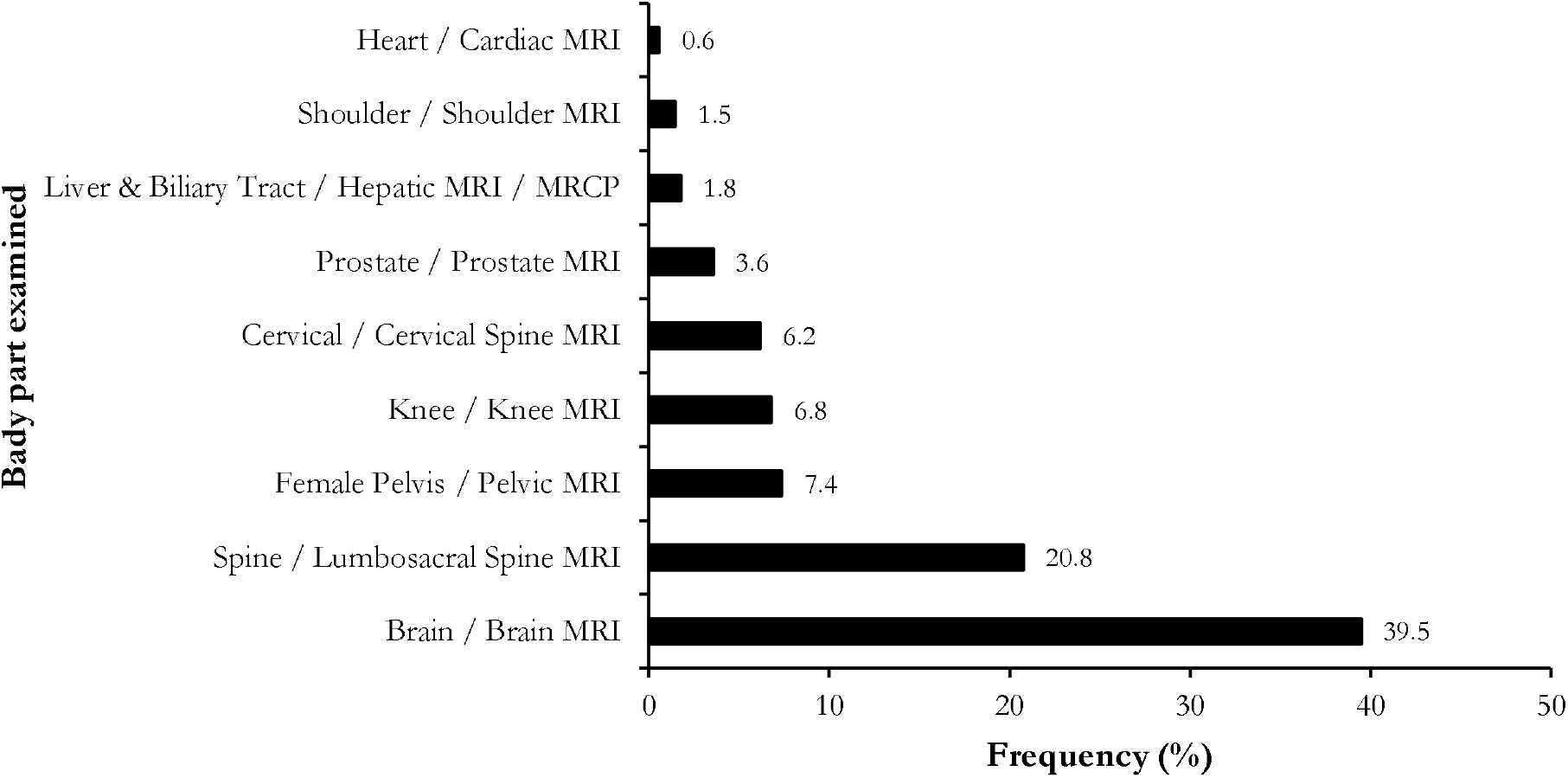
Body part examined by MRI in the studied health facilities, Yaoundé, Cameroon, 2022-2023 (n=337)

The indication for MRI provided was a clinical syndrome or clinical diagnosis in 46.6% of cases; no indication was provided in 0.3% of cases (Fig 4).

**Fig 4.**
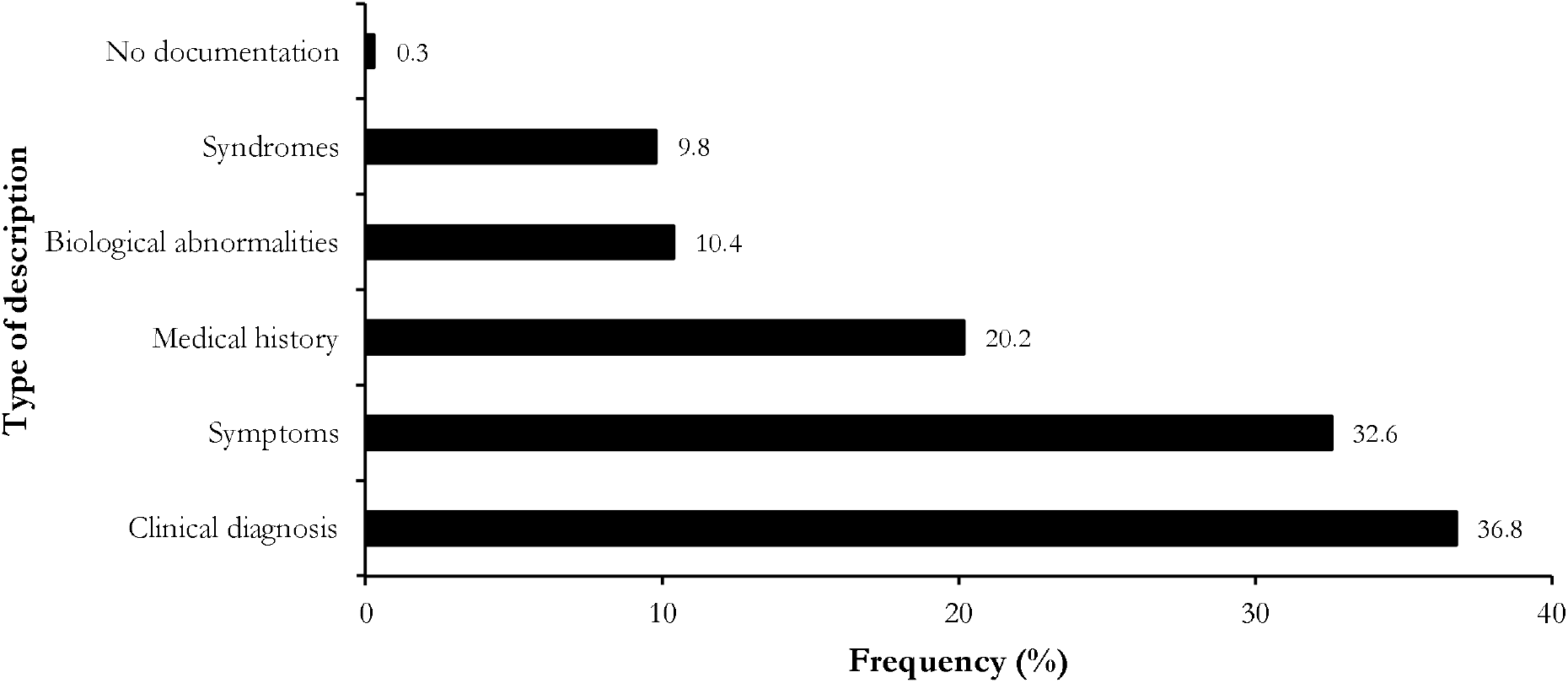
Type of patients’ clinical information provided by the MRI prescribers, Yaoundé, Cameroon, 2022-2023 (n=337)

The purpose of the examination was not mentioned in 56.7% of cases (Table 2).

**Table 2.** MRI demand quality assessment in Yaoundé, Cameroon, 2022-2023 (n=337)

| Variable | Count (n) | Frequency (%) |
| --- | --- | --- |
| **Compatibility of examination requested and information supplied** |  |  |
| Compatible | 326 | 96.7 |
| Non-compatible | 11 | 3.3 |
| Non-analyzable | 0 | 0 |
| **Anatomical region or organ to be examined** |  |  |
| Mentioned | 337 | 100 |
| Not mentioned | 0 | 0 |
| **Purpose of examination or research question** |  |  |
| Mentioned | 146 | 43.3 |
| Not mentioned | 191 | 56.7 |

The examination was relevant in 78.3% of cases and indicated as first-line treatment in 56.1% of cases (Table 3).

**Table 3.** Assessment of MRI prescription relevance, Yaoundé, Cameroon, 2022-2023 (n=337)

| Variable | Count (n) | Frequency (%) |
| --- | --- | --- |
| **Examination relevance** | 264 | 78.3 |
| **Type of examination indication** |  |  |
| Indicated in 1st intention | 189 | 56.1 |
| Indicated in 2nd intention | 47 | 13.9 |
| Not initially indicated | 55 | 16.3 |
| Specialized for the quality of the applicant | 33 | 9.8 |
| Not indicated | 12 | 3.6 |
| Not analyzable | 1 | 0.3 |
| **Applicant qualification** | 331 | 98.2 |

The group of pathologies sought was non-specific at 53.4%, followed by neoplasia at 36.2% (Table 4).

**Table 4.** Breakdown of requests by disease group sought, Yaoundé, Cameroon, 2022-2023 (n=337)

| Variables | Count (n) | Frequency (%) |
| --- | --- | --- |
| **Pathologies searched for** |  |  |
| Non-specific | 180 | 53,4 |
| Malignant neoplasia | 80 | 23,7 |
| Benign neoplasia | 42 | 12,5 |
| Inflammatory/infectious | 34 | 10,1 |
| Vascular | 27 | 8,0 |
| Traumatic | 21 | 6,2 |
| Degenerative | 20 | 5,9 |
| Congenital | 8 | 2,4 |
| Metabolic/ endocrine | 5 | 1,5 |
| **Other pathology** |  |  |
| Foreign Body | 1 | 0,3 |
| Obstructive | 1 | 0,3 |

No socio-demographic data was associated with the relevance of MRI prescription. The relevance of the MRI request was significantly 4 to 5 times higher in the group of requests with clinical syndrome as the indication (OR = 4.7; 95% CI = 1.1 - 20.2; *p* = 0.022). It was significantly 7 to 8 times higher in the group of requests with suspected vascular pathology as the indication (OR = 7.8; 95% CI = 1.1 - 58.9; *p* = 0.014) and significantly 2 times higher in the group of requests with suspected malignant neoplasia as the indication (OR = 2.2; 95% CI = 1.1 - 4.6; *p* = 0.023). The relevance of the MRI request was significantly reduced by 50% (OR = 0.5; 95% CI = 0.3 - 0.9; p = 0.026) in the group of requests with a clinical diagnosis as the indication, and by 40% (OR = 0.6; 95% CI = 0.3 - 0.9; *p* = 0.034) in the group of requests whose indication sought a non-specific pathology (Table 5).

**Table 5.** Univariate analysis of the association between pathologies assessed and MRI prescription relevance, Yaoundé, Cameroon, 2022-2023 (n=337)

| Factor | Total | Relevance<br>n (%) |  | OR | 95% CI Limits |  | p-value |
| --- | --- | --- | --- | --- | --- | --- | --- |
|  |  | Yes | No |  | Lower | Upper |  |
| Age (in years) |  |  |  |  |  |  |  |
| <21 | 31 | 27 (87,1) | 4 (12,9) | 1,7 | 0,5 | 5,5 | 0,367 |
| 21 – 30 | 25 | 23 (92) | 2 (8) | 2 ,9 | 0,6 | 13,4 | 0,171 |
| 30 – 40 | 56 | 45 (80,4) | 11 (19,6) | 1 | 0,4 | 2,3 | 0,933 |
| 40 – 50 | 66 | 46 (69,7) | 20 (30,3) | 0,6 | 0,3 | 1,2 | 0,146 |
| 50 – 60 | 65 | 48 (73,8) | 17 (26,2) | 0,7 | 0,3 | 1,5 | 0,380 |
| 60+ | 94 | 75 (79,8) | 19 (20,2) | - | - | - | - |
| Sexe |  |  |  |  |  |  |  |
| Male | 163 | 123 (75,5) | 40 (24,5) | 0,7 | 0,4 | 1,2 | 0,214 |
| Female | 174 | 141 (81) | 33 (19) | - | - | - | - |
| Health insurance | 69 | 52 (75,4) | 17 (24,6) | 0,8 | 0,4 | 1,5 | 0,501 |
| Pathology |  |  |  |  |  |  |  |
| Traumatic | 21 | 15 (71.4) | 6 (28.6) | 0.6 | 0.2 | 1.8 | 0.427 |
| Vascular | 27 | 26 (96.3) | 1 (3.7) | 7.8 | 1.1 | 58.9 | 0.014 |
| Congenital | 8 | 8 (100) | 0 (0) | NA |  |  |  |
| Degenerative | 20 | 15 (75) | 5 (25) | 0.8 | 0.3 | 2.3 | 0.709 |
| Inflammatory/infectious | 34 | 30 (88.2) | 4 (11.8) | 2.2 | 0.7 | 6.4 | 0.187 |
| Metabolic/endocrine | 5 | 4 (80) | 1 (20) | 1.1 | 0.1 | 10.1 | 1 |
| Benign neoplasia | 42 | 36 (85.7) | 6 (14.3) | 1.7 | 0.7 | 4.3 | 0.215 |
| Malignant neoplasia | 80 | 70 (87.5) | 10 (12.5) | 2.2 | 1.1 | 4.6 | 0.023 |
| Non-specific | 180 | 133 (73.9) | 47 (26.1) | 0.6 | 0.3 | 0.9 | 0.034 |
| Types of information on the request |  |  |  |  |  |  |  |
| Clinical diagnosis | 124 | 89 (71,8) | 35 (28,2) | 0,5 | 0,3 | 0,9 | 0,026 |
| Syndrome | 33 | 31 (93,9) | 2 (6,1) | 4,7 | 1,1 | 20,2 | 0,022 |
| Symptom | 110 | 82 (74,5) | 28 (25,5) | 0,7 | 0,4 | 1,2 | 0,239 |
| Biological abnormalities | 35 | 30 (85,7) | 5 (14,3) | 1,7 | 0,6 | 4,6 | 0,263 |
| Past history | 68 | 60 (88,2) | 8 (11,8) | 2,4 | 1,1 | 5,2 | 0,027 |

OR: Odds Ratio

After multivariate analysis, only the presence of vascular pathology significantly increased the relevance of the request for MRI by eight times (aOR = 8.4; 95% CI = 1.1 - 68.2; *p* = 0.045) (Table 6).

**Table 6.** Multivariate analysis of the association between pathologies assessed and MRI prescription relevance, Yaoundé, Cameroon, 2022-2023 (n=337)

| Parameter | aOR | 95% CI Limits |  | *p*-value |
| --- | --- | --- | --- | --- |
|  |  | Lower | Upper |  |
| Type of information provided |  |  |  |  |
| Clinical diagnosis | 0,6 | 0,3 | 1 | 0,054 |
| Syndrome | 3,6 | 0,8 | 16 | 0,091 |
| Pathology |  |  |  |  |
| Vascular | 8,4 | 1,1 | 68,2 | 0,045 |
| Malignant neoplasia | 2,2 | 0,9 | 5,5 | 0,078 |
| Non-specific | 0,9 | 0,4 | 1,8 | 0,794 |

aOR: *adjusted Odds Ratio*

## Discussion

One of the most difficult decisions for a doctor is to determine the best diagnostic modality and the best time to request it. This can lead to confusion, especially when there is no set of valid criteria. The aim of this study was to investigate the socio-demographic profile of users, and the indication and appropriateness of MRI use by private and public health facilities in sub-Saharan Africa. Following an analytical cross-sectional study with prospective data collection, we registered 337 patients from whom the data that will be discussed in the following lines were collected and analyzed.

The median age of patients was 48 years and the age group over 60 years was the most represented in our series at 27.9%. It should be noted that this result is close to those found in Ghana and Iran [2,8]. There was a predominance of females at 51.6%, i.e. a sex ratio of 0.93, in contrast to the work of Mensah *et al.* in Ghana and that of Gomez-Garcia *et al.* in Spain, where there was a slight predominance of males at 50.6% and 52%. These similarities may be because the study sites were general hospitals that receive patients of all ages and clinical conditions [2,9]. Cerebral MRI was the most frequently performed examination, and 53.4% of patients had an indication for a non-specific pathology group. No socio-demographic data was associated with the relevance of MRI prescription. Adding syndrome as a type of description was associated with a four-fold increase in the relevance of MRI (OR = 4.7; 95% CI = 1.1 - 20.2); *p* = 0.022). Similarly, vascular pathology and malignant neoplasia were significantly associated with a seven-times (OR = 7.8; 95% CI = 1.1 - 58.9; *p* = 0.014) and two-times (OR = 2.2; 95% CI = 1.1 - 4.6; *p* = 0.034) increase in the relevance of MRI respectively. However, when the information on the request was the clinical diagnosis or when the pathology group sought was non-specific, the relevance of the request for examination was significantly reduced by 50% (OR = 0.5; 95% CI = 0.3 - 0.9; *p* = 0.026) and 40% (OR = 0.6; 95% CI = 0.3 - 0.9; *p* = 0.034) respectively. After multivariate analysis, only the presence of vascular pathology significantly increased the relevance of the MRI request by eight times (aOR = 8.4; 95% CI = 1.1 - 68.2; *p* = 0.045).

The request for an examination was deemed relevant in 78.3% of cases and indicated as the first line of defense in 56.1% of cases. However, it was not initially indicated in 16.3% of cases. Lower rates of relevance, at 75.6% and 71.5% respectively, were found in Iran and Canada, favored by the different characteristics of the healthcare systems and coverage by health insurance [8,10]. As a result, a review of decision-making algorithms in the case of additional examinations and improved synergy between clinical doctors and radiologists would optimize patient management and avoid an unnecessary increase in the cost of care. Another explanation would be that the study sites in both ours and theirs were teaching hospitals in which specialists and residents requested diagnostic procedures for therapeutic and educational purposes. A higher rate of inappropriate requests for MRI may be the result of the training process of less experienced doctors [8].

After bivariate statistical analysis, there was no association between socio-demographic profile and the relevance of the examination. These results are comparable to the work of Mensah et al [2], justified by the fact that in our context, optimal patient management is not conditioned by age or socioeconomic status.

The relevance of MRI was reduced by 50% and 40% respectively when the information was the clinical diagnosis and when the pathology group sought was non-specific. Thus, there would be no point in requesting an MRI if the patient’s diagnosis is strongly suspected clinically, or if we don’t really know what we’re looking for. However, a nuance could be made in the context of an extension assessment of a pathology already known in the patient. In our series, the presence of malignant neoplasia doubled the relevance of the examination.

This study also evaluated the clinical conditions on the request forms and the relevance of MRI and showed a more than four-fold increase in the relevance of MRI. This could be because more information on the use of technology has become available to clinicians, increasing the accuracy of its use [8].

In multivariate analysis, after binary logistic regression, the presence of vascular pathology significantly and independently increased the relevance of MRI. Tissue characterization using extracellular volume fraction and T1 mapping has made cardiac MRI invaluable in the diagnostic work-up of infiltrative cardiomyopathy, often negating the need for invasive endomyocardial biopsies. Stress cardiac MRI can also be performed, providing structural information and serving as an ischemic assessment and myocardial viability study.

## Limitations

The main limitation of this study is the selection bias that favors public training to the detriment of private training. However, this study is the first in our context, and the small number of radiology centers with MRI makes our results generalizable to the entire population.

## Conclusion

Most MRI examinations are requested by specialist doctors, with brain and lumbar spine MRIs dominating the requests, while subjects aged 60 and over are the most represented, despite a median patient age of 48. People with health insurance account for 1/5 of patients. Nearly 4/5 of requests were relevant; this level of relevance was increased when the indication provided was a vascular syndrome or pathology and reduced when the indication provided was a clinical diagnosis or non-specific research.

## Data Availability

All data produced in the present work are contained in the manuscript

## Abbreviations

aOR: adjusted Odds Ration

OR: Odds Ratio

CI: Confidence Interval

MRI: Magnetic Resonance Imaging

SPSS: Statistical Package for the Social Sciences

## Declarations

### Authors’ Contribution

Study design & conception: GRPN and BN; Data collection and processing: GRPN; Data analysis, visualization and interpretation: GRPN; Drafting of original manuscript: GRPN, FZLC; Critical revision of the manuscript: FZLC, BM, JTA and JRMT. Final approval of the manuscript: All authors.

### Ethical Approval Statement

The institutional Review Board (IRB) of the Faculty of Medicine and Biomedical Sciences of the University of Yaoundé 1 approved the protocol and an ethical clearance was issued. All methods were performed in accordance with the relevant guidelines of the Helsinki declaration.

### Consent for publication

Not applicable.

### Availability of data and materials

All data generated or analyzed during this study are included in this published article.

